# Immunogenicity of SARS-CoV-2 Vaccine in Dialysis

**DOI:** 10.1101/2021.04.08.21254779

**Authors:** Eduardo Lacson, Christos P. Argyropoulos, Harold J. Manley, Gideon Aweh, Andrew I. Chin, Loay H. Salman, Caroline M. Hsu, Doug S. Johnson, Daniel E. Weiner

## Abstract

**Background:** Patients receiving maintenance dialysis represent a high risk, immune-compromised population with 15-25% COVID mortality rate who were unrepresented in clinical trials evaluated for mRNA vaccines’ emergency use authorization.

**Method:** All patients receiving maintenance dialysis that received two doses of SARS-CoV-2 mRNA vaccines with antibody test results drawn ≥14 days after the second dose, as documented in the electronic health record through March 18, 2021 were included. We report seroresponse based on levels of immunoglobulin-G against the receptor binding domain of the S1 subunit of SARS-CoV-2 spike antigen (seropositive ≥2) using FDA-approved semi-quantitative chemiluminescent assay (ADVIA Centaur® XP/XPT COV2G).

**Results:** Among 186 dialysis patients from 32 clinics in 8 states tested 23±8 days after receiving 2 vaccine doses, mean age was 68±12 years, with 47% women, 21% Black, 26% residents in long-term care facilities and 97% undergoing in-center hemodialysis. Overall seropositive rate was 165/186 (88.7%) with 70% at maximum titer and with no significant difference in seropositivity between BNT162b2/Pfizer (N=148) and mRNA-1273/Moderna (N=18) vaccines (88.1% vs. 94.4%, p=0.42). Among patients with COVID-19 history, seropositive rate was 38/38 (100%) with 97% at maximum titer.

**Conclusion:** Most patients receiving maintenance dialysis were seropositive after two doses of BNT162b2/Pfizer or mRNA-1273/Moderna vaccine. Early evidence suggests that vaccinated dialysis patients with prior COVID-19 develop robust antibody response. These results support an equitable and aggressive vaccination strategy for eligible dialysis patients, regardless of age, sex, race, ethnicity, or disability, to prevent the extremely high morbidity and mortality associated with COVID-19 in this high risk population.

**Significance:** In this retrospective observational evaluation of SARS-CoV-2 mRNA vaccine response defined by detectable levels of immunoglobulin-G against the receptor binding domain of the S1 subunit of SARS-CoV-2 spike antigen of ≥2 in serum of patients receiving maintenance dialysis, 165/186 (88.7%) were found to be seropositive (with 70% at maximum titer) at least 14 days after completing the second dose. No significant differences were observed by race or other subgroup or by vaccine manufacturer. Therefore, an equitable and aggressive vaccination strategy for all eligible maintenance dialysis patients, regardless of age, sex, race, ethnicity, or disability, is warranted to prevent the extremely high morbidity and mortality associated with COVID-19 in this high risk population.

---

Nearly 550,000 people in the United States (US) receive maintenance dialysis and prevalence estimates approach 3 million worldwide.^1,2^ Patients receiving maintenance dialysis (henceforth “dialysis patients”), particularly those receiving hemodialysis, comprise a population of vulnerable individuals who cannot self-isolate, have a high incidence of Severe Acute Respiratory Syndrome Coronavirus 2 (SARS-CoV-2) infection and have Coronavirus Disease 2019 (COVID-19)-associated mortality of 15 to 25%,^3-5^ making effective vaccination a priority. Despite this heightened risk and prior reports of diminished response to vaccines versus other viruses such as hepatitis-B and influenza,^6-8^ the immune response to SARS-CoV-2 vaccination among dialysis patients is unknown due to insufficient trial data. Accordingly, we performed a quality improvement evaluation of multiple Dialysis Clinic, Inc. (DCI) dialysis clinics in the US assessing antibody response following administration of SARS-CoV-2 messenger RNA (mRNA) vaccines.

## Methods

### Study Setting

DCI implemented a standard clinical protocol for systematic measurement of SARS-CoV-2 spike-antibody immunoglobulin-G (SAb-IgG) that physicians could activate for vaccination in the outpatient dialysis clinic, similar to that for hepatitis-B post-vaccination testing.^6^ As access to the SARS-CoV-2 mRNA vaccines for dialysis patients varied among and within states, they were obtained from nursing homes, hospitals, locally designated centers and pharmacies, and only in few dialysis clinics. Individual physicians ordered post-vaccination SAb-IgG titers based on patient accessibility and clinical interest. All vaccinations were verified and recorded in the DCI electronic health record (EHR). This interim report includes a retrospective evaluation of all patients receiving 2 doses of either BNT162b2/Pfizer or mRNA-1273/Moderna vaccine per manufacturer’s recommendation across 32 DCI clinics in 8 states with SAb-IgG measured ≥14 days after the second dose, documented as of March 18, 2021.

### SARS-CoV-2 Spike Antibody Assay

DCI Lab measured SAb-IgG against the receptor binding domain (RBD) of the S1 subunit of SARS-CoV-2 spike antigen using US-FDA-approved chemiluminescent assay (ADVIA Centaur® XP/XPT COV2G).^9, 10^ This semi-quantitative assay has a range between 0 and ≥20, with a positive threshold of ≥2. S1-RBD-antibodies are relevant to vaccines incorporating this immune-dominant region to elicit neutralizing (and therefore likely protective) antibodies in vaccinated individuals.^11^

### Electronic Health Record

Vaccination dates, SAb-IgG results, demographic and clinical data including age, sex, race/ethnicity, body mass index, dialysis parameters (i.e. vintage, modality, access, adequacy), albumin, number and types of comorbidities, use of immunomodulatory agents, history of transplantation or COVID-19 diagnosis, and other vaccines administered or hospitalization within 14 days of vaccination were obtained from the DCI-EHR. This retrospective evaluation is a DCI quality improvement evaluation under Western Institutional Review Board (WIRB) exemption. Statistical analyses were performed using SAS v9.4.

## Results

Among 186 dialysis patients receiving 2 doses of vaccine, mean age was 68±12 years, with 47% women, 21% Black, 26% residents in long-term care facilities and 97% undergoing in-center hemodialysis (**Table 1**). SAb-IgG was positive (≥2) in 165/186 (88.7%) with no significant difference between BNT162b2/Pfizer (N=148) and mRNA-1273/Moderna (N=18) vaccines (88.1% vs. 94.4%, p=0.42). The distribution of titer levels drawn over 23±8 days after the 2^nd^ dose is shown in **Figure 1**. Non-responders had mean SAb-IgG titer of 0.4±0.2 vs. 18.2±4.3 among responders (p<0.0001).

**Table 1.** Patient characteristics presented as mean ± standard deviation or number (percent).

| **Demographics** | **All Patients**<br>**(N=186)** | **Responders**<br>**(N=165)** | **Non-Responders**<br>**(N=21)** |
| --- | --- | --- | --- |
| **Age (years)** | 67.9 ± 12.2 | 67.7 ± 12.4 | 70.0 ± 10.7 |
| ≥ 65 years | 123 (66.1) | 107 (64.8) | 16 (76.2) |
| **Female** | 88 (47.3) | **73 (44.2)** | **15 (71.4)<sup>a</sup>** |
| **Race** |  |  |  |
| White | 73 (39.3) | 62 (37.6) | 11 (52.4) |
| Black | 39 (21.0) | 36 (21.8) | 3 (14.3) |
| Native American | 33 (17.7) | 31 (18.8) | 2 (9.5) |
| Asian/Pacific Islander | 19 (10.2) | 18 (10.9) | 1 (4.8) |
| Other/Unknown | 22 (11.8) | 18 (10.9) | 4 (19.1) |
| **Hispanic** | 15 (8.1) | 14 (8.5) | 1 (4.8) |
| **Vintage (months)** | 58.1 ± 54.3 | **60.6 ± 54.2** | **38.7 ± 52.2<sup>a</sup>** |
| **Body Mass Index (kg/m<sup>2</sup>)** | 28.7 ± 7.1 | 28.7 ± 6.9 | 29.2 ± 8.8 |
| **Long Term Care Facility** | 48 (25.8) | 41 (24.9) | 7 (33.3) |
| **Home Dialysis** | 6 (3.2) | 5 (3.0) | 1 (4.8) |
| **Dialysis Access** |  |  |  |
| Fistula | 129 (69.4) | 117 (70.9) | 12 (60.0) |
| Graft | 29 (15.6) | 26 (15.8) | 3 (15.0) |
| Central Venous Catheter | 23 (12.4) | 18 (10.9) | 5 (25.0) |
| Peritoneal Dialysis Catheter | 5 (2.7) | 4 (2.4) | 1 (4.8) |
| **Adequate Dialysis Dose<sup>c</sup>** | 174 (93.6) | 155 (93.9) | 19 (90.5) |
| **Serum Albumin (g/dl)** | 3.8 ± 0.4 | 3.8 ± 0.4 | 3.5 ± 0.6 |
| **Other Vaccines within 14 days** | 20 (10.8) | **15 (9.1)** | **5 (23.8)<sup>a</sup>** |
| Pneumonia | 4 (2.2) | 3 (1.8) | 1 (4.8) |
| Hepatitis B | 17 (9.1) | 13 (7.9) | 4 (19.1) |
| **Potential Immunosuppression** | 30 (18.2) | **30 (18.2)** | **8 (38.1)<sup>a</sup>** |
| Immune-modulating Meds | 29 (15.6) | 23 (13.9) | 6 (28.6) |
| Prior Transplant | 10 (5.4) | 8 (4.9) | 2 (9.5) |
| Immunodeficiency Disorder | 10 (5.4) | 6 (3.6) | 4 (19.1) |
| **Hospitalization within 14 days** | 34 (18.3) | **24 (14.6)** | **10 (47.6)<sup>b</sup>** |
| **Disability<sup>d</sup>** | 17 (9.1) | 17 (10.3) | 0 (0.0) |
| **Tobacco Use** | 22 (11.8) | 21 (12.7) | 1 (4.8) |
| **Alcohol Abuse Disorder** | 14 (7.5) | 14 (8.5) | 0 (0.0) |
| **Drug Abuse Disorder** | 8 (4.3) | 6 (3.6) | 2 (9.5) |
| **COVID-19 History** | 38 (20.4) | **38 (23.0)** | **0 (0.0)<sup>a</sup>** |
| **Number of Comorbidities** | 3.2 ± 1.9 | 3.2 ± 1.9 | 3.6 ± 1.7 |
| Diabetes Mellitus | 129 (69.4) | 113 (68.5) | 16 (76.2) |
| Hypertension | 152 (81.7) | 134 (81.2) | 18 (85.7) |
| **Congestive Heart Failure** | 37 (19.9) | **27 (16.4)** | **10 (47.6)<sup>b</sup>** |
| COPD<sup>e</sup> | 34 (18.3) | 30 (18.2) | 4 (19.1) |
| Stroke/Cerebrovascular Disorder | 23 (12.4) | 19 (11.5) | 4 (19.1) |
| Peripheral Vascular Disease | 25 (13.4) | 22 (13.3) | 3 (14.3) |
| Thyroid Disorder | 36 (19.4) | 30(18.2) | 6 (28.6) |
| History of Cancer | 18 (9.7) | 14 (8.5) | 4 (19.1) |

<sup>a</sup> P-value <0.05; <sup>b</sup> P-value <0.01; <sup>c</sup> Adequate dialysis defined by hemodialysis dose spKt/V≥1.2 or peritoneal dialysis dose weekly Kt/V≥1.7; <sup>d</sup> Amputee, wheelchair use or inability to perform activities of daily living; <sup>e</sup> COPD=Chronic Obstructive Pulmonary Disease. Note: Comparisons between responders with non-responders utilized Chi-Square or Analysis of Variance for categorical variables and T-test / Kruskal-Wallis/Mann-Whitney test for normal/non-normally distributed continuous variables.

**Figure 1.**
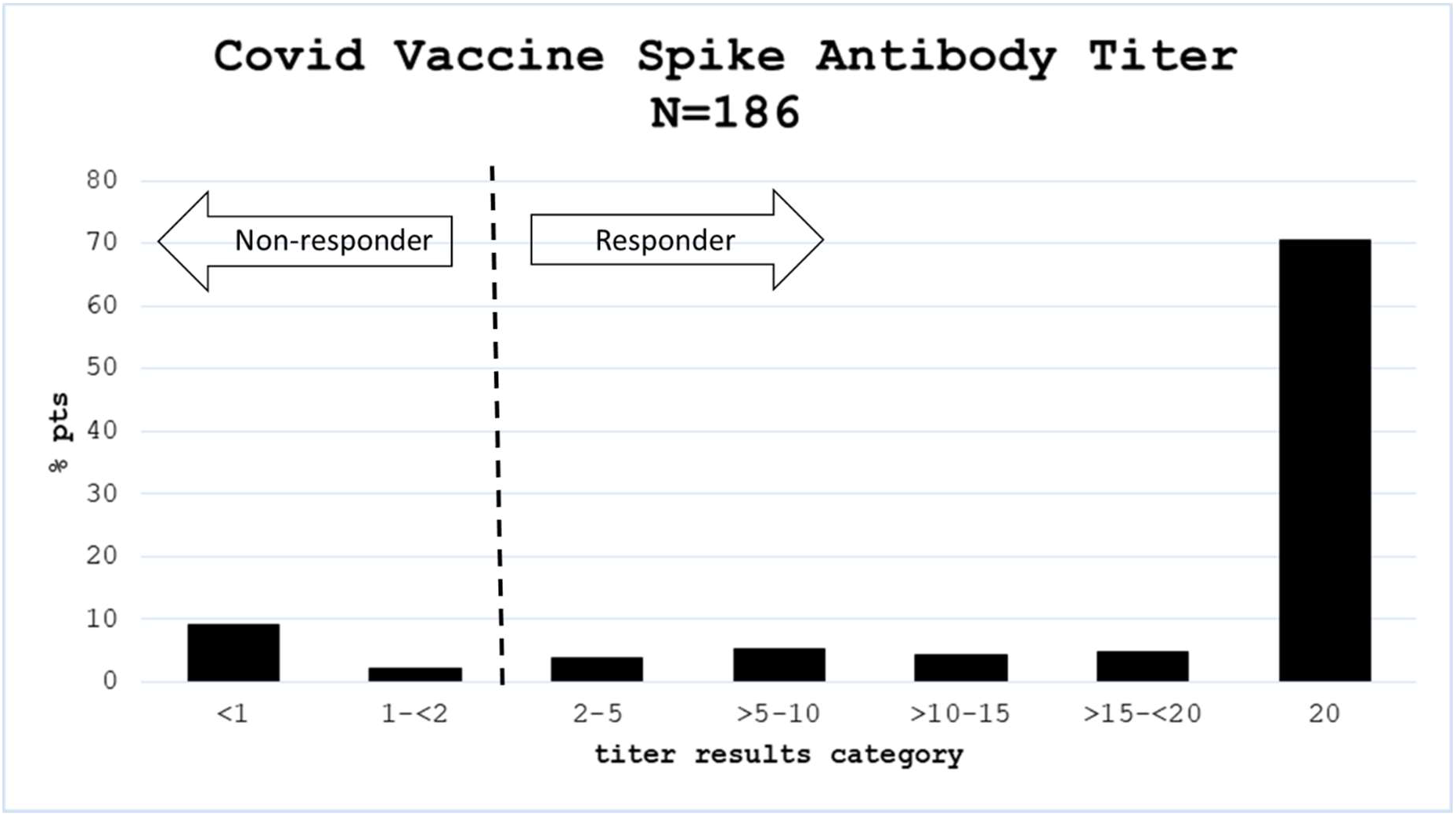
Distribution of spike antibody IgG titers for SARS-CoV-2 mRNA vaccinated maintenance dialysis patients (N=186) drawn over 23±8 days after the second dose.

Among 148 patients without known COVID-19, 127 (85.8%) were seropositive following vaccination. All 38 patients with COVID-19 diagnosis were seropositive, irrespective of vaccine type, with 37/38 at maximum limit of the assay. In 36 patients, COVID-19 diagnosis occurred 181±118 days prior to completing vaccination. The remaining two patients were long-term care facility residents each exposed to a roommate, and were tested while asymptomatic at 1 and 5 days after their second vaccine dose, respectively (with SAb-IgG titers obtained 20 days and 23 days after vaccination, respectively). Upon review, the latter patient had recent gastrointestinal upset and constipation but neither was hospitalized.

Seroresponse occurred in 73 of 88 women versus 92 of 98 men (83% vs. 94%, p=0.02). Results of univariate analysis are indicated in **Table 1**. Non-responders were newer to dialysis and more likely to have other vaccines administered or be hospitalized within 14 days of SARS-CoV-2 vaccination, have potential immunosuppressed state related to immune-modulating drugs, prior transplant, and/or immune-deficiency disorders, and congestive heart failure (CHF). A summary of the clinical characteristics of non-responders is provided in **Table 2**.

**Table 2.**
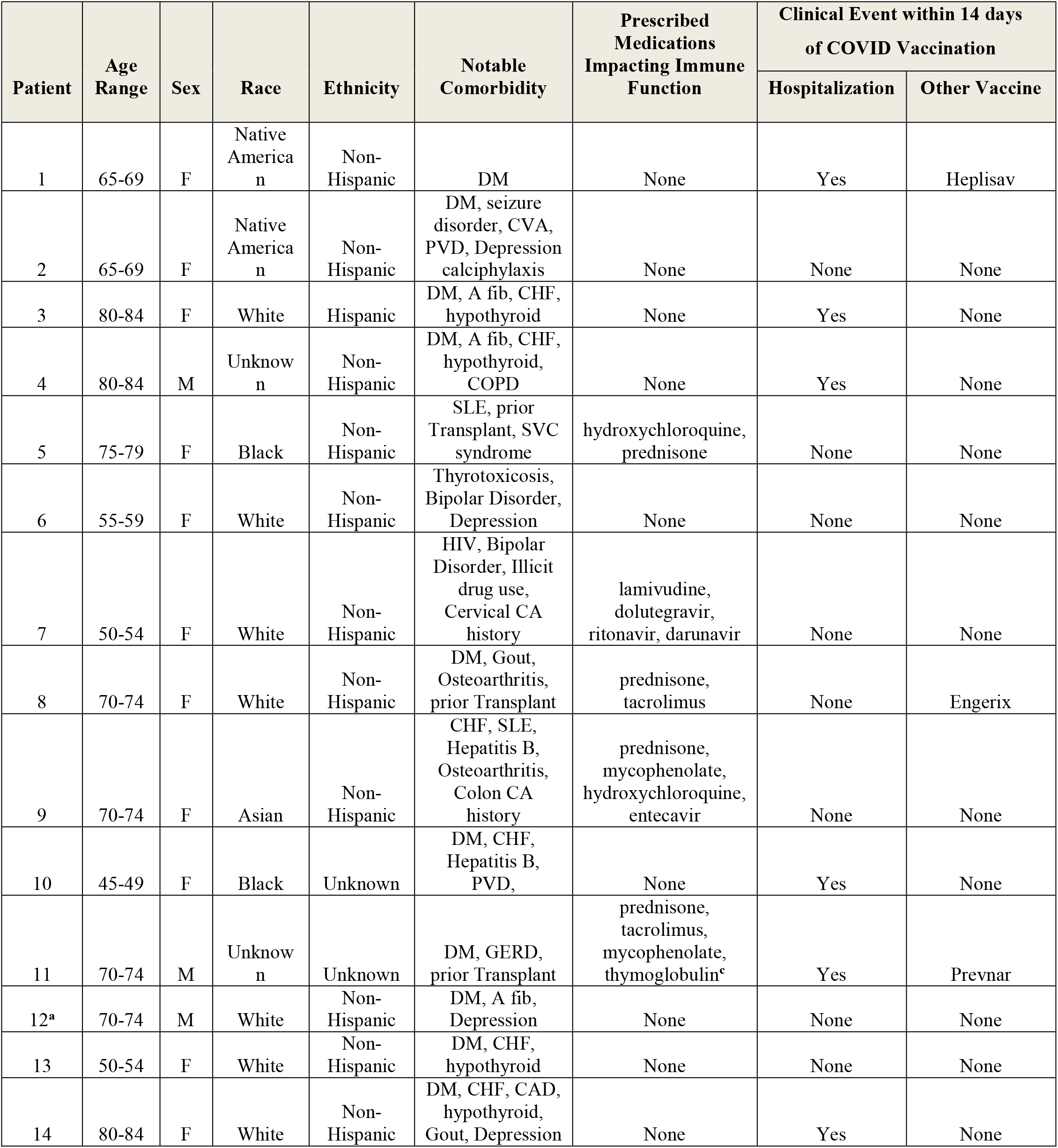
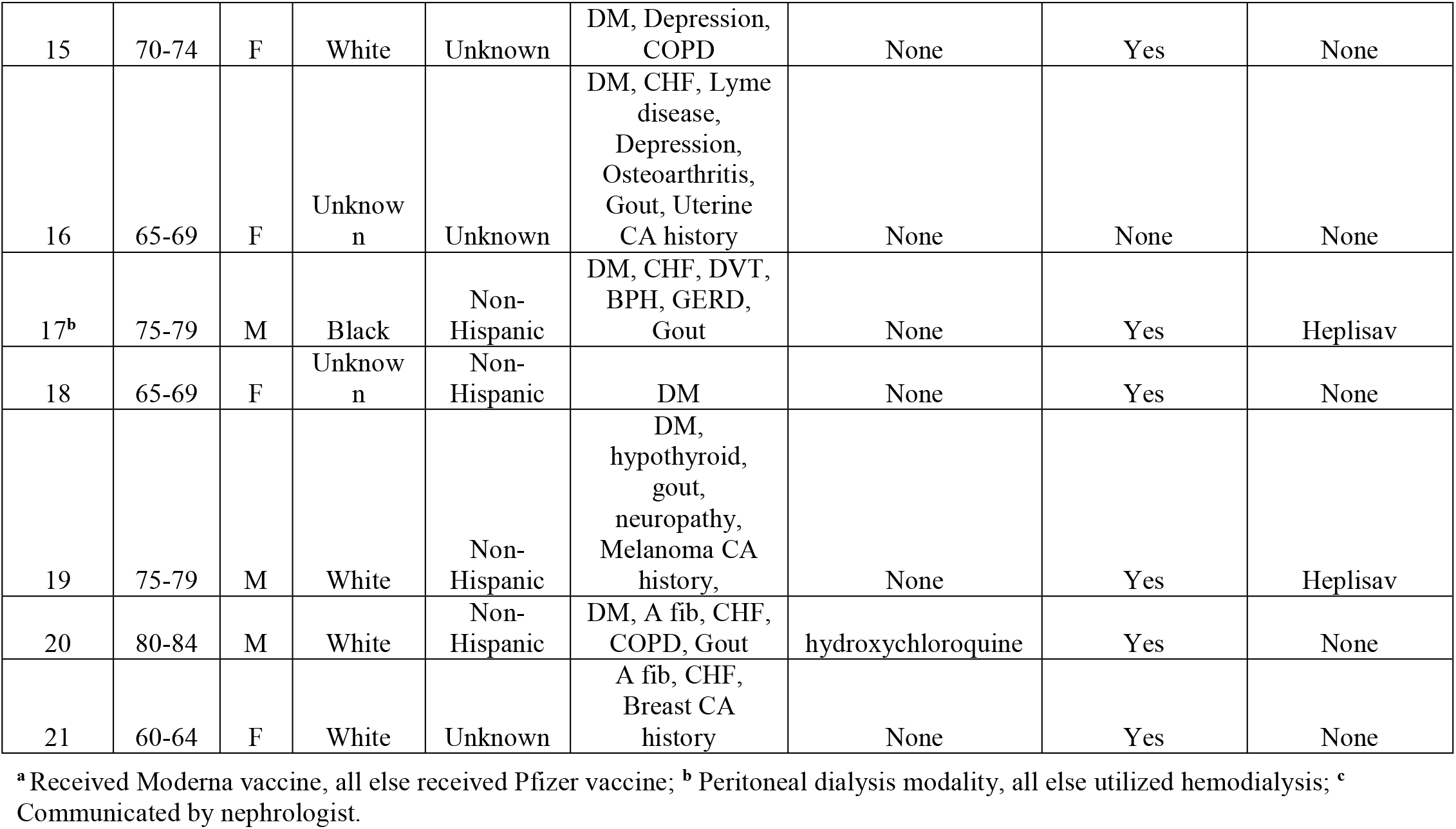
Detailed summary of characteristics of SARS-CoV-2 mRNA vaccine non-responders.

## Discussion

Among maintenance dialysis patients in the US, 88.7% achieved SAb-IgG >2, measured ≥14 days after two doses of SARS-CoV-2 mRNA vaccine. Near universal seroconversion rates occurred in clinical trials utilized for these vaccines’ emergency use authorization, but notably did not include patients on dialysis. Lower seroconversion rate was not unexpected, given the widespread immune dysfunction observed in dialysis-dependent patients. These immune alterations included skewed Th1/Th2 responses, impaired function of the professional antigen presenting cells (APC), and susceptibility of B-cells to apoptosis.^12^ The combined deficits in T-cell, APC and B-cell functions render dialysis patients less likely to both seroconvert and maintain protective titers over time compared to individuals without kidney disease,^13^ previously observed with pneumococcal, hepatitis-B and H1N1 influenza vaccines.^7,8,14^

Factors associated with poor seroconversion in our cohort include female sex, younger vintage, potential immunosuppression from diseases, transplant or medications, CHF and co-vaccination and hospitalization during the peri-vaccination period. A recent meta-analysis suggests women receiving dialysis respond to vaccines equally to men, although differences in seroconversion rates by sex were noted in some hepatitis-B series.^7^ However, we caution that some of these associations, including that for women, may not be sustained as we accumulate more data. Of note, potential immunosuppressed states and acute hospitalization during the vaccine series have face validity to impact immune response, particularly in the setting of uremic inflammation.^7,12,15^

Considering the devastating impact of COVID-19 on dialysis patients compared to the potentially dramatic benefit of vaccination on severity of illness,^3-5^ it is imperative to explore strategies to monitor and maximize seroconversion. Such knowledge may be useful for patient education and dialogue to reinforce preventive measures. Strategies to improve seroconversion may include additional or higher vaccine doses. Monitoring longitudinal SAb-IgG could inform potential use of booster doses, similar to algorithms used for hepatitis-B vaccination. Such tracking is a component of the quality program at DCI and will be critical in informing the development of alternative vaccination regimens.

A strength of the current report is coverage of multiple areas of the country with a diverse dialysis population. Although limited by small sample size, the data appear sufficient to emphasize that seroconversion should be expected for the majority of maintenance dialysis patients. Additional limitations include an observational design that may have led to residual confounding and selection biases, heavily influenced by accessibility and availability of vaccines. Future studies could also evaluate T-cell responses given that specific clinical implications of serology tests including the SAb-IgG we utilized, need further elucidation.

In conclusion, the vast majority of maintenance dialysis patients responded with IgG spike antibody titers to a complete series of both SARS-CoV-2 mRNA BNT162b2 and mRNA-1273 vaccines. Further study is needed to determine duration of the seroprotection as well as the ideal approach to non-responders, including whether they should receive a booster dose. Early evidence suggests that vaccinated dialysis patients with prior COVID-19 develop robust antibody response. Our findings support an equitable and aggressive vaccination strategy for all eligible maintenance dialysis patients regardless of age, sex, race, ethnicity, or disability, to prevent the extremely high morbidity and mortality associated with COVID-19 in this high risk population.

## Data Availability

n/a

## Acknowledgment

We are grateful for the assistance from DCI Medical Directors and Clinic Managers caring for our patients who worked to have our patients vaccinated against SARS-CoV-2 and tirelessly documented the information in the EHR. We further thank the nephrologists who have implemented DCI clinical and quality improvement protocols and care recommendations, including the SARS-CoV-2 seroresponse testing algorithm. We also thank Beth Kammer from DCI Laboratories, Vlad Ladik and Brian Tinger from DCI IT-Analytics Department, Karen Majchrzak with DCI Research and Senior/Area Operations Directors who supported this quality improvement project. Dr. Hsu is funded by T32-DK007777.

